# An International Consensus Definition for Contextual Factors: Findings from a Nominal Group Technique

**DOI:** 10.1101/2022.12.16.22283573

**Authors:** Chad E Cook, Antoine Bailliard, Jennifer Bent, Joel Bialosky, Elisa Carlino, Luana Colloca, Jorge E Esteves, Dave Newell, Alvisa Palese, William R. Reed, Jennifer Plumb Vilardaga, Giacomo Rossettini

## Abstract

Emerging literature suggests contextual factors are important components of therapeutic encounters and may substantially influence clinical outcomes of a treatment intervention. At present, a single consensus definition of contextual factors, which is universal across all health-related conditions is lacking. The objective of this study was to create a consensus definition of contextual factors to better refine this concept for clinicians and researchers. The study used a multi-stage virtual Nominal Group Technique (vNGT) to create and rank contextual factor definitions. Nominal group techniques are a form of consensus-based research, and are beneficial for identifying problems, exploring solutions and establishing priorities. The 10 international vNGT participants had a variety of clinical backgrounds and research specializations and were all specialists in contextual factors research. The initial stages of the vNGT resulted in the creation of 14 independent contextual factor definitions. After a prolonged discussion period, the initial definitions were heavily modified, and 12 final definitions were rank ordered by the vNGT participants from first to last. A sixth round was used to identify a final consensus, which reflected the complexity of contextual factors and included three primary domains: 1) an overall definition; 2) qualifiers that serve as examples of the key areas of the definition; and 3) how contextual factors may influence clinical outcomes. Our consensus definition of contextual factors seeks to improve the understanding and communication between clinicians and researchers. These are especially important in recognizing their potential role in moderating/mediating clinical outcomes.

## Introduction

Tools such as patient reported outcome measures (PROMs), physical performance measures, and patient experience measures, are used to measure a patient’s health outcomes [1], and are influenced by a number of internal (within the person) and external (outside the person) factors. These factors may include comorbidities [2], cognition and mood [2], socioeconomic and social status [3,4], and care timing and provider specialization [5-7]. Targeted treatment/interventions may also influence outcomes but are commonly moderated and/or mediated by factors such as expectations [8,9], the patient-clinician relationship [10], legal status [11], workers compensation [11], social risk variables [3,4], common factors [12], and natural history [13]. These factors influence disparate individuals differently; consequently, understanding the role that interventions contribute toward patient outcomes becomes challenging.

The ecological landscape in which the clinical encounter occurs, sometimes referred to as therapeutic context, constituting a range of factors increasingly referred to as contextual factors, can also markedly moderate or mediate outcomes [14]. Although increasingly well studied, contextual factors/effects are defined differently across a majority of studies [10, 15-21]. Definitions vary and have included sociodemographic variables [15], person-related factors (race, age, patient beliefs and characteristics) [15], and physical and social environments [16]. At a micro-level, contextual factors have been defined by seemingly disparate terms such as therapeutic alliance [10], one’s role in the environment [17], treatment characteristics [18], healthcare processes [19], placebo or nocebo effects [20], government agencies [17], and cultural beliefs. Occasionally, at a macro-level, they are described as confounders or effect modifiers that are not an outcome of the study, but need to be recognized (and measured) [16,21].

Recently, through a multi-step process, the Outcome Measures in Rheumatology (OMERACT) initiative created a consensus definition for contextual factors [21]. The principal goal of OMERACT was to identify contextual factors that were relevant for clinical trials. Initially, OMERACT defined a contextual factor as a “*variable that is not an outcome of the study, but needs to be recognized (and measured) to understand the study results. This includes potential confounders and effect modifiers*” [22]. Through semi-structured interviews and Delphi research, the OMERACT group further qualified contextual factor types (relevant for clinical trials) as: 1) effect modifying (those that modify the treatment effect); 2) outcome influencing (those that predict the prognosis and may confound results); and 3) measurement affecting (those that influence measurement properties such as reliability and validity).

The OMERACT’s broad definition is useful for understanding results in a clinical trial, in that it exists within a more historic paradigm that seeks to remove effects rather than enhance them. In this role it fails to resolve some of the confusion associated with the multitudes of ways contextual factors are presently defined (specifically, whether internal and external domains are potentially contextual factors), does not include qualifiers to improve one’s understanding, and provides no guidance as to how clinicians may identify contextual factors within clinical encounters in order to enhance positive and minimize negative effects. Subsequently, the objective of this study was to create a consensus definition of contextual factors to better encapsulate this concept both to guide clinicians in clinical scenarios as well as broaden definitions for researchers. This study used a virtual nominal group technique (vNGT) [23], and included researchers and research clinicians from multiple professions who specialized in the study of contextual effects research. Similar to the OMERACT group, we endeavored to identify a consensus definition that reflects the complexity of contextual factors and describe how contextual factors may influence clinical outcomes, but were also interested in a more detailed set of qualifiers that serve as examples of the key areas of the definition.

## Methods

### Study Design

The mixed methods study used a vNGT [23]. The vNGT was performed in October of 2022. Nominal group techniques are beneficial for identifying problems, exploring solutions and establishing priorities, and encourages contributions from all participants and treats each person equally [23]. The Institutional Review Board of Duke University, Durham, North Carolina, USA, approved the study (ro00111522-INIT-1.0).

### Nominal Group Technique Participants

Optimal NGT participants are stated as five to nine individuals [23], but values may vary. NGT participants were identified by their expertise in contextual factors and/or by their targeted clinical background or specialization and invited to participate in the vNGT.

### Study Procedure

Participants were provided with pre-work prior to the vNGT. Each individual was provided with an article [23] that outlined the vNGT processes and were asked to consider early development of their own versions of a definition for contextual factors.

During the virtual session, a five-stage NGT process following the protocol by Potter et al. [23] was used (Figure 1). The virtual session was conducted using Microsoft Zoom (Microsoft Corp, Redmond, WA) and the moderator for the session was a mixed-methods researcher with a contextual factors background and prior experience with vNGT research and moderation.

**Fig. 1.**
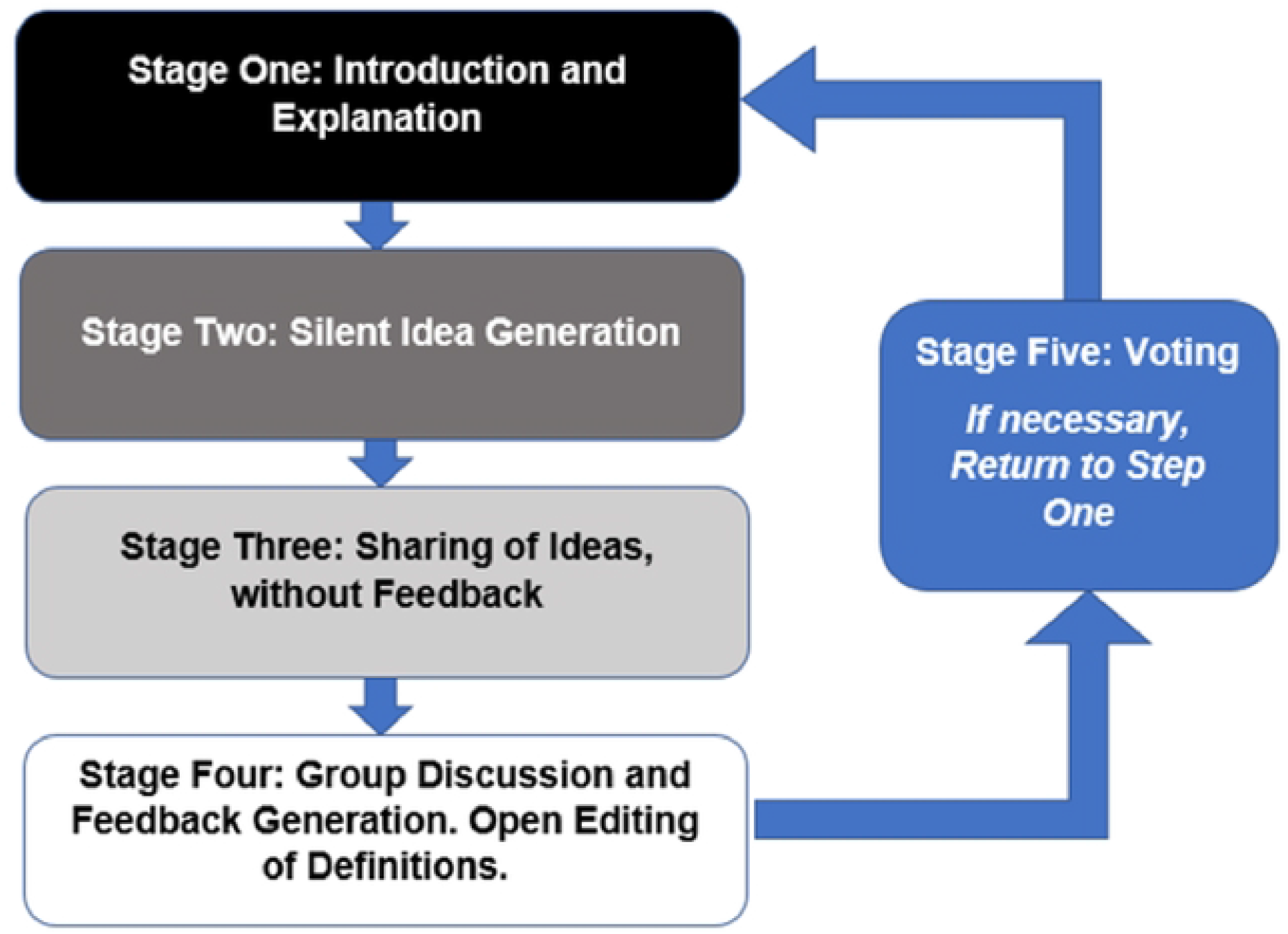
The Five Stage Process of a Virtual Nominal Group Technique according to Potter [23]. <u>Stage one</u> (Introduction and Explanation): An introduction and welcome to all participants with an explanation of the purpose and procedure of the workshop. <u>Stage two</u> (Silent Idea Generation): The question was introduced to the participants: “What is a working definition of contextual factors”? All participants were asked to create a list of ideas that come to mind when considering the question and to place these ideas on a shared Google document. During this stage, all participants were asked not to consult or discuss ideas with each other. A total of 10 minutes was provided for each participant to create their selected definitions. <u>Stage three</u> (Sharing Ideas): During Stage three, each participant introduced their definitions that were recorded on the google documents. This document was shared on the screen so that all participants can see the list in real time. This stage continued in a round robin format until all ideas had been presented. No debate or discussion occurred at this stage. <u>Stage four</u> (Group Discussion): Participants were invited to seek verbal explanation or further details about any ideas that were produced during stage three. The moderator ensured that each person was able to contribute and that all ideas were discussed without spending too long on a single idea. At this stage, participants were able to suggest new items for discussion or combining of items to modify the current list. Each participant “owned” each definition and edited the definition only if they agreed on the change requested. Unique to this vNGT, participants had up to one week to modify or delete their own contributions or request edits to another definition that they did not generate. We elected to provide additional time to edit each person’s definition, since the concept is complex and since there were a variety of definitions presented in Stage two and three, which were further discussed and modified in Stage four. <u>Stage five</u> (Voting): During stage five, and after the week of modifying or deleting their own contributions, vNGT participants were allowed to “rank order” the definitions generated during stage four. Rank ordering was performed using a Qualtrics survey and a “ranking” function. In this survey, each NGT participant ranked all 12 definitions from 1 (top choice) to 12 (lowest choice).

Modifications of a five round NGT are not uncommon and may be warranted when working with complex populations or topics that require maturation before final evaluation [24]. If consensus voting does not identify a clear ranked winner, a sixth round, which includes re-voting on the top ranked choices, can be implemented to assure a true consensus choice [23,24]. Our vNGT used a sixth round of voting to identify a clear consensus definition.

## Results

### Participants Characteristics

The NGT included 10 individuals with clinical/research backgrounds in rehabilitation (chiropractic, osteopathy, physical therapy, or occupational therapy), clinical psychology, medicine, and nursing. Advanced research training included community engagement, molecular biology, nursing science, neurobiology, neuroscience, placebo/nocebo, rehabilitation medicine, and social determinants of health (Table 1). The NGT participants averaged 19 publications on contextual factors, and represented 4 countries across two continents.

**Table 1.** Virtual Nominal Group Technique Voting Participants’ Backgrounds.

| Clinical Background | Research Training and Background | Location | Number of Publications involving Contextual Factors |
| --- | --- | --- | --- |
| Chiropractic | Anatomical Sciences and Neurobiology<br>Neuroscience with a research focus in physiological and pain-related mechanisms | United States | 30 |
| Medical Physician / Neurophysiologist | Neuroscience | United States | 10 |
| None | Molecular Biology, Musculoskeletal Health | UK | 9 |
| Nursing | Nursing Science | Italy | 15 |
| Occupational Therapy | Community-based mental health service delivery models, sensory processing and participation | United States | 19 |
| Osteopathy | Cognitive Science | Malta | 10 |
| Physiotherapist | Placebo and Nocebo effects associated with contextual factors | Italy | 18 |
| Physiotherapist | Rehabilitation Science | United States | 21 |
| Psychologist | Neuroscience | Italy | 34 |
| Psychologist | Behavioral Intervention, Development and implementation, and Mediators and Moderators of Intervention Effects | United States | 21 |

### Findings of the Nominal Group Technique

Stage two generated 14 definitions of contextual factors (Table 2). Seven vNGT participants submitted one definition, whereas two participants submitted two definitions and one submitted three. Consistent domains included internal and external factors, which influenced outcomes associated with any of the treatments provided. Stage three refined the definitions through audience (clinicians and researchers) discussion and the need for a single consensus definition including qualifiers that help define the definition and how contextual factors may influence outcomes.

**Table 2.**
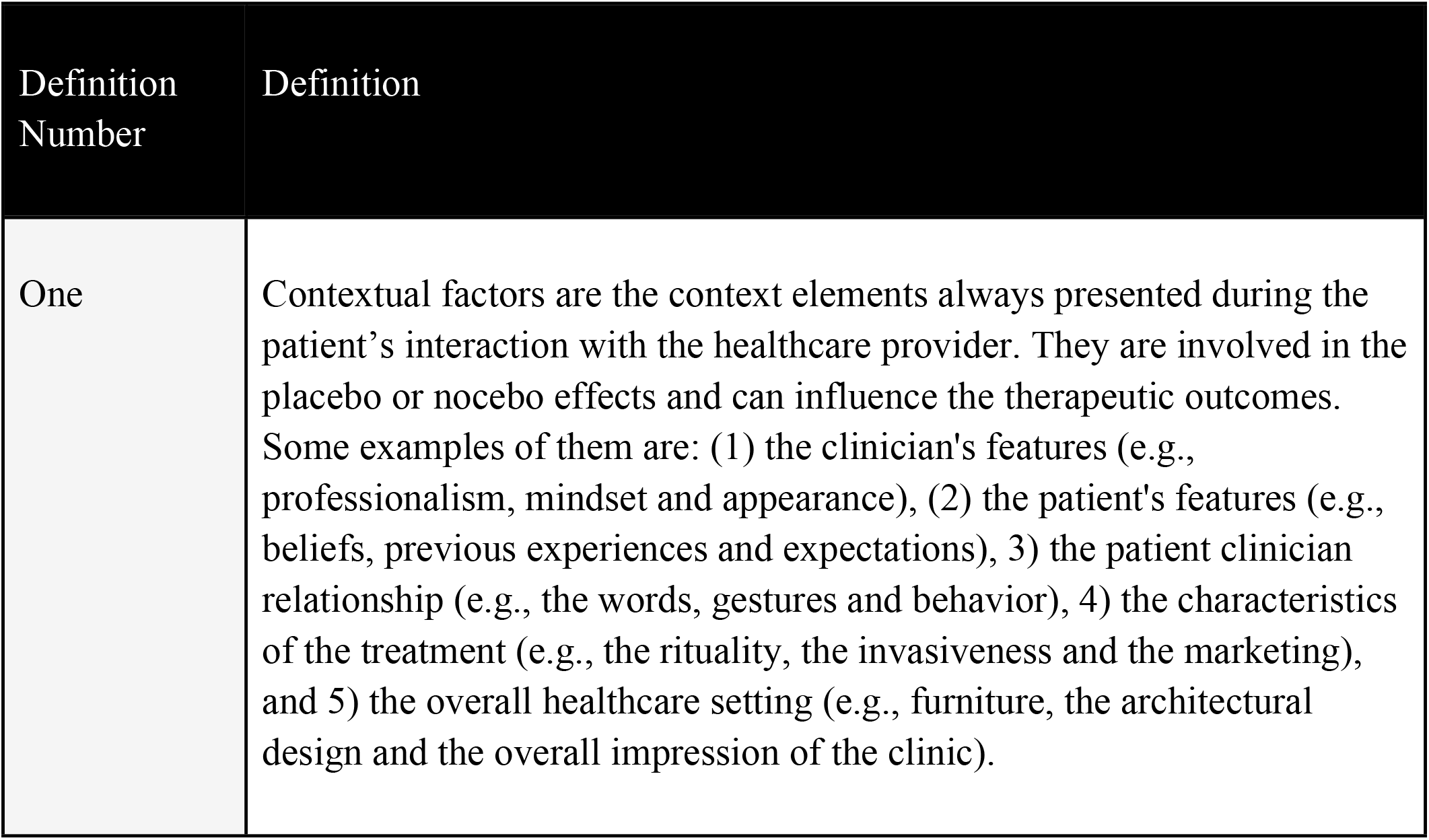
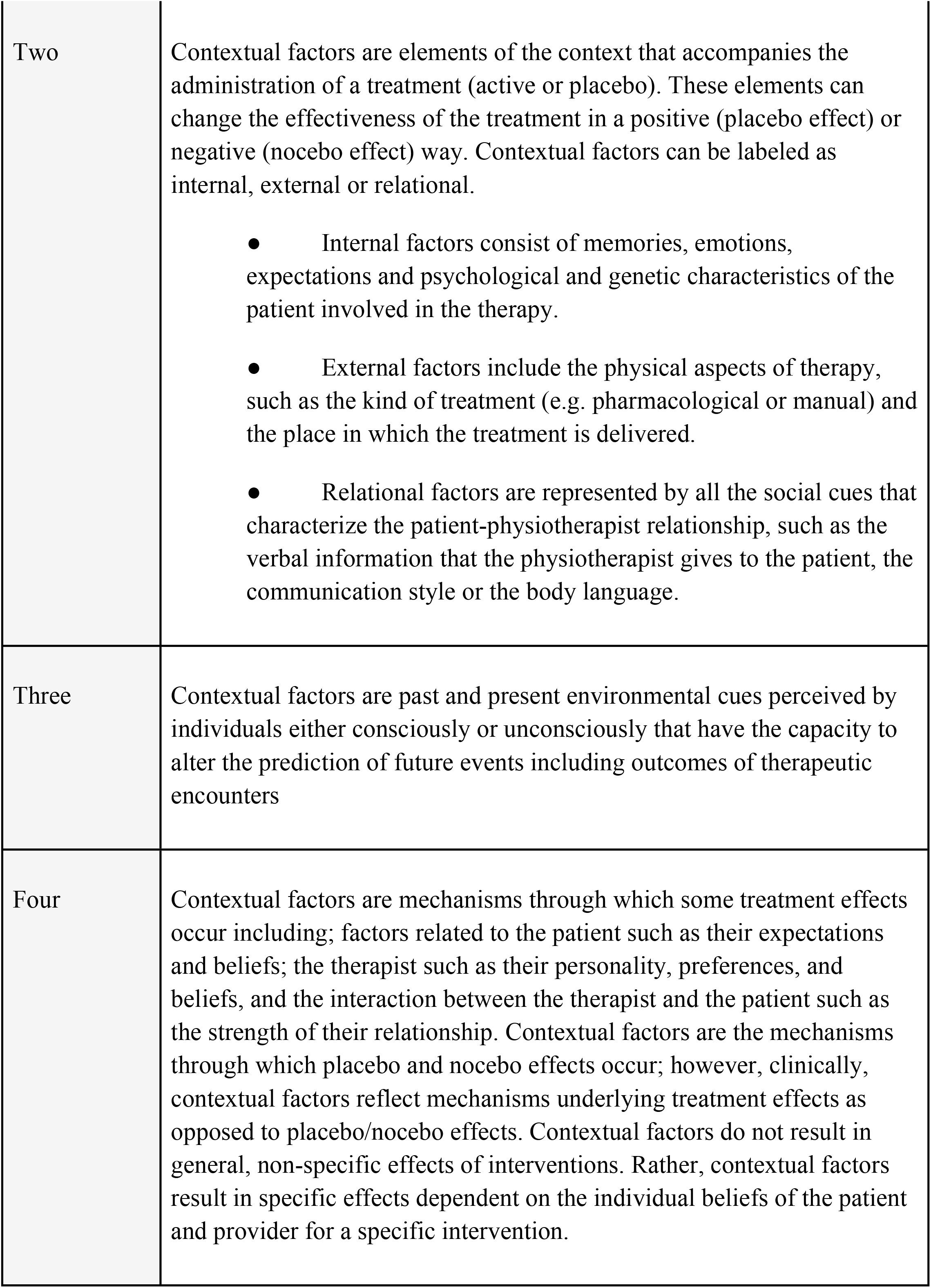
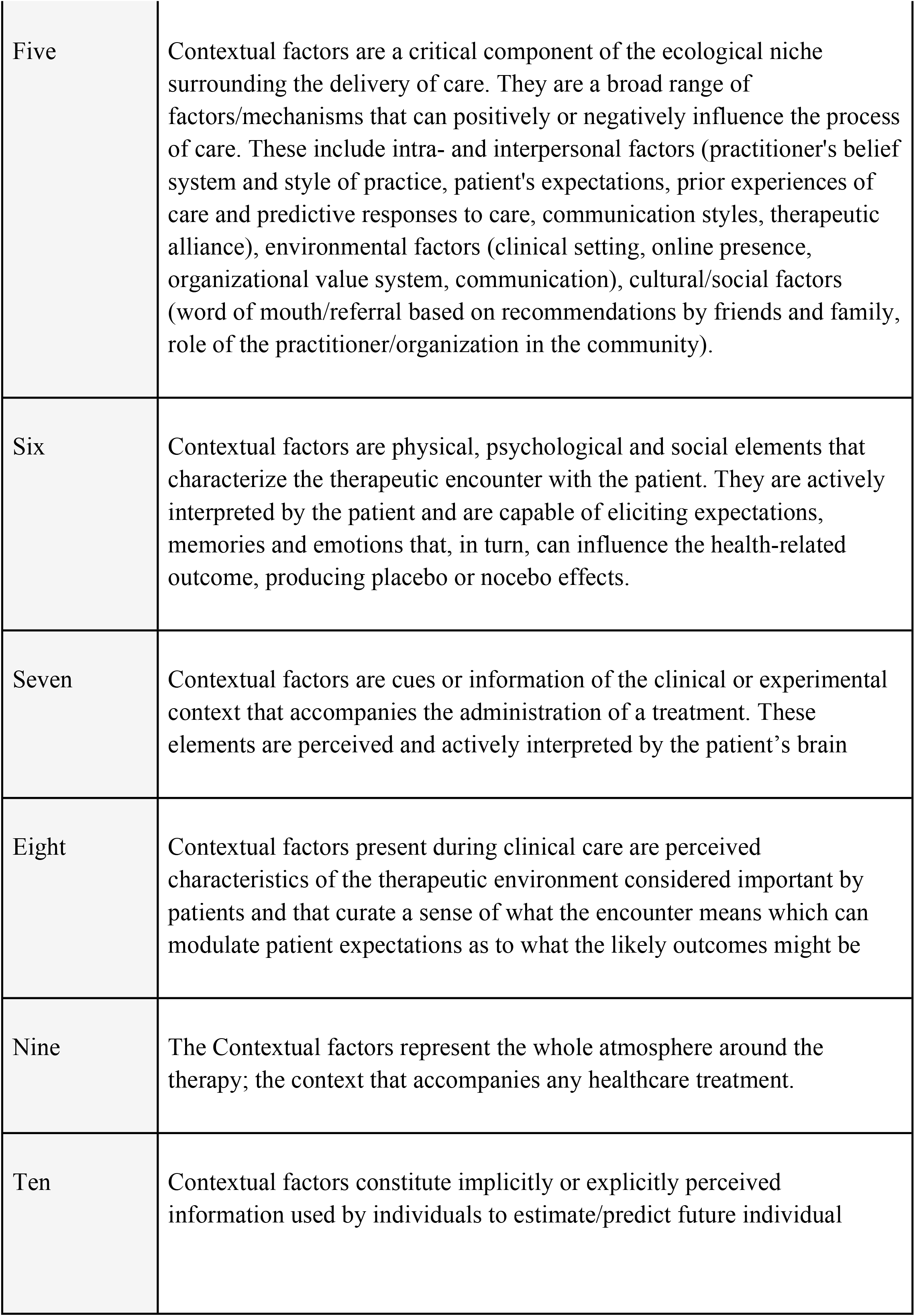
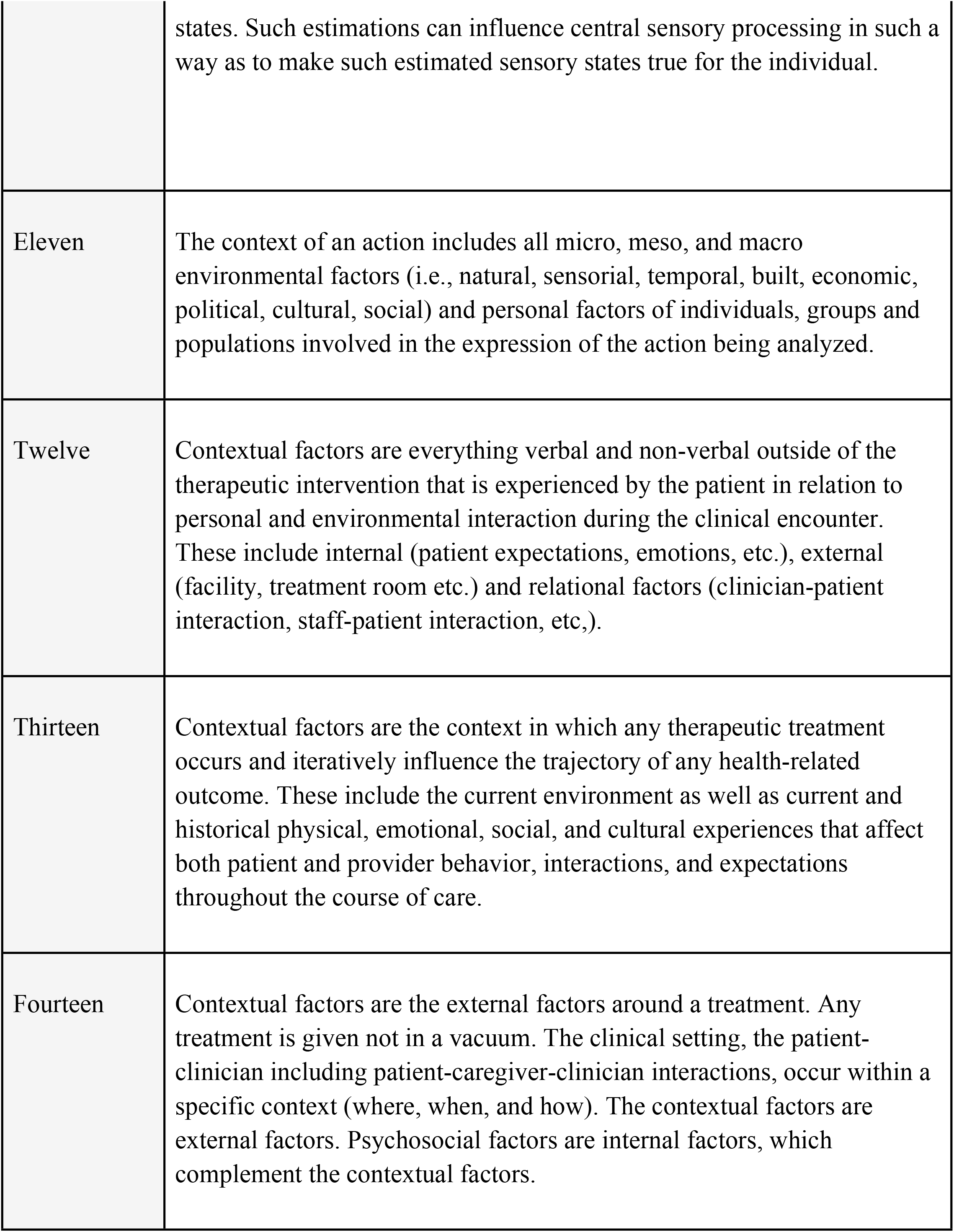
Initial Contextual Factor Definitions (Upon Completion of Stage Two).

At the end of Stage four (consolidation of ideas), there were 12 definitions that were rank ordered (Table 3). Three definitions were clearly ranked higher than (Table 4) the remaining nine with the majority (80%) of the vNGT selecting these choices as one of the top three selections.

**Table 3.** Modified Contextual Factor Definitions (Upon Completion of Stage Four)

| Definition Number | Definition |
| --- | --- |
| One | Contextual factors (CFs) are components of the therapeutic encounter whereby interventions, medications, pharmacological and nonpharmacological treatments are given. CFs encompass the patient and provider personal (i.e., race/ethnicity and expectations), historical (i.e., clinical history and prior experiences), cultural (i.e., social norms, spirituality/religion and power differentials), environmental (i.e., settings and rituals), physical (i.e., sensorial perception and clinical procedures), and rhetorical (i.e., verbal and non-verbal elements of communication) dimensions around the therapeutic encounter and the patient-clinician interaction influencing moderators/mediators of therapeutic mechanisms and the response to any interventions/treatments and ultimately, the overall clinical outcomes. CFs can change the effectiveness of the treatment in a positive (placebo effect) or negative (nocebo effect) way. |
| Two | Contextual factors (CFs) are components of the therapeutic encounter whereby interventions, medications, pharmacological and nonpharmacological treatments are given. CFs encompass the patient and provider personal (e.g., race/ethnicity, genetic variables, expectations, values and preference), historical (e.g., clinical history, prior experiences), cultural (e.g., social norms, spirituality/religion and power differentials), environmental (e.g., settings and rituals), physical (e.g., sensorial perception, clinical examination and modalities in which the therapy is delivered), and rhetorical (e.g., verbal and non-verbal communication) dimensions around the therapeutic encounter and the patient-clinician interaction influencing moderators/mediators of therapeutic mechanisms and the response to any interventions/treatments and ultimately, the overall clinical outcomes. CFs can change the effectiveness of the treatment in a positive (placebo effect) or negative (nocebo effect) way. |
| Three | Contextual factors are mechanisms through which some treatment effects occur including; factors related to the patient such as their expectations and beliefs; the therapist such as their personality, preferences, and beliefs, and the interaction between the therapist and the patient such as the strength of their relationship. Contextual factors are the mechanisms through which placebo and nocebo effects occur; however, clinically, contextual factors reflect mechanisms underlying treatment effects as opposed to placebo/nocebo effects. Contextual factors do not result in general, non-specific effects of interventions. Rather, contextual factors result in specific effects dependent on the individual beliefs of the patient and provider for a specific intervention. |
| Four | Contextual factors are a critical component of the ecological therapeutic niche. They are a broad range of factors that can positively or negatively influence the process of care. These include intra- and interpersonal factors (practitioner's belief system and style of practice, patient's expectations, prior experiences of care and predictive responses to care, communication styles, therapeutic alliance), environmental factors (clinical setting, online presence, organizational value system, communication), cultural/social factors (word of mouth/referral based on recommendations by friends and family, role of the practitioner/organization in the community). |
| Five | Contextual factors are physical, psychological and social elements that characterize the therapeutic encounter with the patient. They are actively interpreted by the patient and are capable of eliciting expectations, memories and emotions that, in turn, can influence the health-related outcome, producing placebo or nocebo effects. |
| Six | Contextual factors are cues or information of the clinical or experimental context that accompanies the administration of a treatment. These elements are perceived and actively interpreted by the patient's brain. |
| Seven | Contextual factors represent the whole atmosphere around the therapy; the context that accompanies any healthcare treatment. |
| Eight | Contextual factors (CFs) are components of all therapeutic encounters and may constitute the entirety of the perceived effects of the intervention itself or be additive to effects of interventions such as pharmacological and nonpharmacological treatments. CFs are perceived cues that affect both the patient and practitioner and can arise from experiences and immediate dynamics within the encounter, or a combination of both. CFs fall into broad categories that can include patient characteristics, practitioner characteristics, treatment characteristics, characteristics of the dynamic between the patient and practitioner and characteristics of the setting within which the encounter is being delivered. CFs can be complexly interwoven in the patients and practitioners experience so as to influence what patients and practitioners expect the outcome of the encounter to be. Through such conscious and unconscious expectations, involving a range of specific neurological pathways, CFs can directly impact (both positively and negatively) symptoms and characteristics associated with the presenting condition. The proportion of clinical effects observed associated with CFs can vary from large to small depending on the characteristics of the patient, practitioner, condition and intervention. |
| Nine | Contextual factors are integral components of a therapeutic encounter and can include environmental factors (e.g., natural, sensorial, temporal, built, economic, political, cultural, social) and personal factors of all individuals involved (e.g., physical, mental, social, cultural) in the therapeutic encounter. Some contextual factors are modifiable and can be targeted in intervention to effect change to personal factors. |
| Ten | Contextual factors are everything verbal and non-verbal outside of the therapeutic intervention that is experienced by the patient in relation to personal and environmental interaction during the clinical encounter. These include internal (e.g. patient expectations, emotions, cultural), external (e.g. facility ambience, environment) and relational (e.g. clinician/staff-patient interaction, social, physical, historical) factors that impact moderators/mediators of therapeutic outcomes. |
| Eleven | Contextual factors are moderating/mediating components of the therapeutic encounter that influence the trajectory of a health-related outcome. These include the current therapeutic environment as well as current and historical physical, emotional, social, and cultural experiences that affect both patient and provider behavior, interactions, and expectations throughout the course of care. |
| Twelve | Contextual factors are components of the therapeutic encounter whereby interventions, medications, pharmacological and nonpharmacological treatments are given. Contextual factors encompass the patient and provider personal (e.g., race/ethnicity, expectations, values and preference), historical (e.g., clinical history, prior experiences), cultural (e.g., social norms, spirituality/religion and power differentials), environmental (e.g., settings and rituals), physical (e.g., sensorial perception, and clinical examination), and rhetorical (e.g., verbal and non-verbal communication) dimensions around the therapeutic encounter and the patient-clinician interaction influencing moderators/mediators of therapeutic mechanisms and the response to any interventions/treatments and ultimately, the overall clinical outcomes. |

**Table 4.** Top Three Ranked Contextual Factor Definitions (Upon Completion of Stage Five).

| Rank<br>Order | Definition | Average Score<br>/ Median |
| --- | --- | --- |
| First | Contextual factors are components of the therapeutic encounter whereby interventions, medications, pharmacological and nonpharmacological treatments are given. Contextual factors encompass the patient and provider personal (e.g., race/ethnicity, expectations, values and preference), historical (e.g., clinical history, prior experiences), cultural (e.g., social norms, spirituality/religion and power differentials), environmental (e.g., settings and rituals), physical (e.g., sensorial perception, and clinical examination), and rhetorical (e.g., verbal and non-verbal communication) dimensions around the therapeutic encounter and the patient-clinician interaction influencing moderators/mediators of therapeutic mechanisms and the response to any interventions/treatments and ultimately, the overall clinical outcomes. | 3.0 / 2.0 |
| Second | Contextual factors (CFs) are components of all therapeutic encounters and may constitute the entirety of the perceived effects of the intervention itself or be additive to effects of interventions such as pharmacological and nonpharmacological treatments. CFs are perceived cues that affect both the patient and practitioner and can arise from past experiences and immediate dynamics within the encounter, or a combination of both. CFs fall into broad categories that can include patient characteristics, practitioner characteristics, treatment characteristics, characteristics of the dynamic between the patient and practitioner and characteristics of the setting within which the encounter is being delivered. CFs can be complexly interwoven in the patients and practitioners experience so as to influence what patients and practitioners expect the outcome of the encounter to be. Through such conscious and unconscious expectations, involving a range of specific neurological pathways, CFs can directly impact (both positively and negatively) symptoms and characteristics associated with the presenting condition. The proportion of clinical effects observed associated with CFs can vary from large to small depending on the characteristics of the patient, practitioner, condition and intervention. | 3.7 / 3.0 |
| Third | Contextual factors (CFs) are components of the therapeutic encounter whereby interventions, medications, pharmacological and nonpharmacological treatments are given. CFs encompass the patient and provider personal (e.g., race/ethnicity, genetic variables, expectations, values and preference), historical (e.g., clinical history, prior experiences), cultural (e.g., social norms, spirituality/religion and power differentials), environmental (e.g., settings and rituals), physical (e.g., sensorial perception, clinical examination and modalities in which the therapy is delivered), and rhetorical (e.g., verbal and non-verbal communication) dimensions around the therapeutic encounter and the patient-clinician interaction influencing moderators/mediators of therapeutic mechanisms and the response to any interventions/treatments and ultimately, the overall clinical outcomes. CFs can change the effectiveness of the treatment in a positive (placebo effect) or negative (nocebo effect) way. | 3.8 / 3.0 |

These three were similar in content and scope and finished with mean “ranked” scores of 3.0, 3.7 and 3.8 respectively. Following a further poll of the group it was felt that it was necessary to vote again (Round six), but to only include the three aforementioned definitions. Upon re-vote, one clear winner was identified.

#### Final Definition

*Contextual factors (CFs) are components of all therapeutic encounters and may constitute the entirety of the perceived effects of the intervention itself or be additive to effects of interventions such as pharmacological and nonpharmacological treatments. CFs are perceived cues that affect both the patient and practitioner and can arise from previous experiences and immediate dynamics within the encounter, or a combination of both. CFs fall into broad categories that can include patient characteristics, practitioner characteristics, treatment characteristics, characteristics of the dynamic between the patient and practitioner and characteristics of the setting within which the encounter is being delivered. CFs can be complexly interwoven in the patients and practitioners experience so as to influence what patients and practitioners expect the outcome of the encounter to be. Through such conscious and unconscious expectations, involving a range of specific neurological pathways, CFs can directly influence (both positively and negatively) symptoms and characteristics associated with the presenting condition. The proportion of clinical effects observed associated with CFs can vary from large to small depending on the characteristics of the patient, practitioner, condition and intervention*.

## Discussion

The goal of the study was to develop a consensus-derived definition of contextual factors. The study methodology used an vNGT, which is beneficial for identifying problems [25], exploring solutions and establishing priorities, and providing a meaningful and economical method of soliciting contributions from all participants [23]. Our final consensus selection reflects the complexity of a definition of contextual factors and includes: 1) an overall definition, 2) qualifiers that serve as examples of the key areas of the definition and 3) how contextual factors may influence clinical outcomes. We feel this harmonized definition will improve the understanding of contextual factors and will help clinicians recognize their potential role in moderating/mediating these factors to positively impact clinical outcomes. Further, we feel the findings may also improve interpretation of research and deserve additional discussion.

The NGT participants (similar to the OMERACT group [21]) identified the influence of contextual factors as mediators, moderators, or confounding variables and felt that contextual factors included both internal and external factors. These fell into broad categories that included patient characteristics, practitioner characteristics, treatment characteristics, characteristics of the dynamic between the patient and practitioner and characteristics of the setting within which the encounter is being delivered. This suggests that *who* is enrolled in a study, *who* provides care in a study, and *where* that study is performed may influence clinical outcomes. Because of this, studies require careful discussion on these aspects in their methodology and a discussion of their potential to influence outcomes in their results.

Our initial set of 14 definitions identified a number of common elements associated with contextual factors. The biggest differences across initial definitions included whether contextual factors were considered as placebo/nocebo effects, whether the factors were actively or passively perceived (or both), and whether contextual factors were considered moderators of treatment (e.g., age, sex, socioeconomic status), mediators of treatment (e.g., self-efficacy, fear, psychological mood) or both. Discussion during stage three highlighted the inconsistent domains involved in the role of cultural versus political versus power imbalances, whether contextual factors were a measurable mechanism, whether placebo/nocebo effects were a necessity within the definition, if a contextual factor was a “nonspecific” finding, and its role as a prognostic mediator/moderator. Thus, the emerging findings mirror the heterogeneity of conceptual definition and the variability of dimensions associated with contextual factors reported in the literature [14-21]. Our work acknowledged and established an initial synthesis of these complex and important domains, which may in turn be fruitful to consider in future work.

Despite a wide range of clinical backgrounds and research training expertise, we were pleased with the collaborative nature of our vNGT. With appropriate pre-work and judicious use of time [23], we were able to consolidate many disparate initial thoughts to common themes within the two-hour timeframe. When properly employed, consensus agreement methods create structured environments for which experts are prompted to give the best available information, allowing solutions to problems that may remain otherwise unsolved [26]. This requires the process to be deliberately inclusive, participatory, collaborative, and cooperative, with an ultimate goal of a final consensus agreement [27]. A fundamental element of this methodology is that it does not require all participants to agree on all topics (it implies only general agreement) nor does it assure unanimity.

At the end of stage four, vNGT participants were allocated one week to modify their own definitions of contextual factors and then were given a 48-hour window to rank order the final definitions. Eventually, a sixth round was deemed necessary to further separate three competing definitions.During stage four, notable harmonizing occurred across each of the definitions, especially our first goal of obtaining an overall definition. A majority also agreed that contextual factors moderated or mediated clinical outcomes and compared to the initial set of definitions, most included qualifying statements with the definitions as well. The qualifying statements, such as patient and provider personal, historical, cultural, environmental, physical, and rhetorical dimensions around the therapeutic encounter and the patient-clinician interaction, are what separates our definition from that of the OMERACT group [21].

### Limitations

Although this study provided new insights into contextual factors’ definition, some limitations are worth mentioning. Firstly, although we exceeded the recommended panel size of an NGT, we involved a small sample of participants from a restricted number of healthcare fields, possibly leaving others unrepresented (e.g., midwifery, speech therapy, and optometry). Regardless, we ensured adequate representativeness and geodiversity of contextual factors experts in our vNGT by balancing the number of males and females (M: F = 6:4) and including clinicians and clinical researchers throughout the world [28]. Secondly, compared to conducting an in-person NGT, using a virtual Zoom platform could have produced a limited interaction between participants with potentially diverging opinions, thus introducing bias. Nevertheless, the limited time and resource requirements of the vNGT, together with the presence of an experienced moderator, guaranteed a satisfactory quality of the participatory process, considering all participants’ views equally and minimizing any dominant effects [28]. Lastly, we should have compared the definition of contextual factors obtained with our vNGT with other methods (e.g., Delphi, brainstorming) to evaluate their similarities and differences. However, we deliberately used the vNGT because it represents a suitable consensus method to reach an agreement on a single and complex topic among the participants [28].

## Conclusion

Our study, involving a panel of international experts, offered the opportunity to identify a definition of contextual factors, find their qualifiers and understand their impact on the therapeutic outcome. Our findings may help clinicians and researchers embrace the complexity that underlies the construct of contextual factors. We acknowledge different opinions can coexist; we present our definition as a starting point for future studies on the topic.

## Data Availability

All relevant data are within the manuscript and its Supporting Information files.

